# Extracellular vesicle–derived urinary metabolomes show distinctive changes with breast cancer

**DOI:** 10.64898/2026.01.05.26343426

**Authors:** Anuradha U K H Bambarandhage, Nur Aimi Aliah Zainurin, Noura Laziri, Tim Gate, Helen Tench, Manfred Beckman, Helen Phillips, Russell Morphew, Mandana Pennick, Luis A J Mur

## Abstract

**Introduction:** Breast Cancer (BC) remains a significant clinical challenge, and despite well-established screening strategies, new biomarkers could improve BC detection, treatment and management. Urine represents a headily accessible liquid biopsy for diagnosis and extracellular vesicle (EV) transfer of oncogenic proteins, RNAs, and metabolites that promote tumor growth, invasion, metastasis, and immune evasion.

**Aims:** To compare the whole urine and urinary EV metabolomes and identify BC specific metabolite changes.

**Methodology:** Urine samples were collected from four participant groups: breast cancer (BC) patients (n = 42), individuals with breast benign disease (BBD; n = 3), symptom controls (SC; n = 4), and healthy controls (HC; n = 6). EVs were isolated using differential centrifugation, ultrafiltration, and size-exclusion chromatography (SEC), and their morphology was confirmed by transmission electron microscopy (TEM). Metabolites from whole urine and from EVs derived from the same samples were extracted using methanol–water (70:30, v/v) and analyzed by direct-infusion mass spectrometry (DI-MS) in both positive and negative ESI modes. Metabolic features were processed with BinneR and annotated using the HMDB and KEGG databases. Integrated multi-omics analysis of whole-urine and EV-associated metabolomes was performed using the DIABLO framework within the MixOmics package in R platform.

**Results:** DI-MS profiling detected a broad spectrum of metabolites in both whole–urine and EV–derived fractions. Multivariate analyses revealed a clear separation of breast cancer (BC) patients from healthy controls and non–cancer groups in both matrices. Whole EV metabolites with area under the curves (AUC) of > 0.7 included glyceryl phosphoryl derivatives, N-eicosapentaenoyl species, sphinganine-1-phosphate and tetracosahexaenoic acid. EV–enriched metabolites included carnitine, histidine and adenosine monophosphate. DIABLO–based integrative analysis suggested that urinary and EV metabolomes were broadly similar with the discrete putative metabolite biomarkers representing minor, but specific changes with BC.

**Conclusions:** The whole urine and EV metabolomes suggested a small number of metabolite changes that were specific to BC. This could indicate that the urinary EVs describe distinctive aspects of the breast carcinogenic process.

## Introduction

Breast cancer (BC) poses a considerable challenge to global health, and it is the most frequently diagnosed cancer worldwide, with an estimated 2.26 million cases reported in 2020. Malignant neoplasms also represent the most significant global health challenge for women, accounting for an estimated 107.8 million Disability-Adjusted Life Years (DALYs), with BC alone contributing 19.6 million DALYs (Bray et al., 2018, Ferlay et al., 2019) .

In the United Kingdom, more than one-third of BC cases are diagnosed in women aged 70 and above, while fewer than one in five women are diagnosed before reaching the age of 50. Conversely, in less developed nations, over 50% of BC cases occur in women under 50 years of age (Heer et al., 2020). As life expectancy improves in tandem with economic growth in these regions, an increase in the incidence of BC is anticipated (Sung et al., 2021).

Risk factors for BC include age, a family history of breast malignancy, genetic mutation and reproductive history (Łukasiewicz et al., 2021). BC symptoms can include alterations in breast size, shape, or contour, as well as the presence of a mass or lump. The presence of a breast lump represents the most common presenting symptom and carries a relatively high predictive value for malignancy. At its early stages BC can be asymptomatic. Symptoms such as nipple discharge or bleeding and cutaneous changes may also occur and, in some cases, persist for more than a year before diagnosis (Koo et al., 2017; Walker et al., 2014).

BC prevention strategies include promoting breastfeeding, avoiding tobacco use, reducing obesity, limiting alcohol intake, vaccinating against oncogenic infections, and increasing physical activity (Wild et al., 2020; McTiernan et al., 2008). Obesity (BMI >30) increases BC risk due to enhanced oestrogen production by adipose tissue and elevated levels of insulin and insulin-like growth factor-1 (IGF-1), both of which promote cancer cell proliferation (Momenimovahed & Salehiniya, 2019). Excess adipose tissue also contributes to chronic inflammation through the secretion of pro-inflammatory cytokines, leptin, and vascular endothelial growth factor (VEGF), alongside reduced levels of the protective adipokine adiponectin. This inflammatory state activates the nuclear factor kappa-light-chain-enhancer of activated B cells (NF-κB) pathway, which drives tumor cell proliferation, angiogenesis, invasion, and metastasis (Smolarz et al., 2022). Alcohol consumption elevates circulating estrogen levels by impairing hepatic metabolism and enhancing the aromatisation of androgens to estrogens, thereby activating estrogen receptor pathways that support the growth of hormone receptor–positive BC (Smolarz et al., 2022). Tobacco smoke contains numerous carcinogens capable of inducing DNA damage and mutations, contributing to cancers such as lung, oesophageal, bladder, and cervical cancers (Alexandrov et al., 2016; Khani et al., 2018). Combined alcohol and tobacco use further exacerbates BC risk due to their synergistic effects, particularly increased estrogen exposure and heightened carcinogen burden (Loroña et al., 2024).

Apart from these preventive measures, prompt and accurate BC diagnosis substantially enhances treatment outcomes and overall survival. Breast Self-Examination (BSE) and Clinical Breast Examination (CBE) are mainly used for cancer screening. Whilst CBE has a high specificity of 97.11%, its sensitivity is relatively lower at 57.14% (Ratanachaikanont, 2005). Mammograms are routinely utilized for the assessment and identification of breast abnormalities, and clinically, digital mammography (DM) is most often used in the diagnosis of BC. However, breast tumors must be at least several millimeters in size for effective detection.

At the molecular level, changes in the expression of estrogen receptor (ER), progesterone receptor (PR), and human epidermal growth factor receptor 2 (HER2) are routinely assessed in BC. They are associated with poorer prognoses (Asiago et al., 2010). These alterations may be linked to inherited mutations in BRCA1 and BRCA2, which together account for approximately 30–50% of known hereditary BC mutations (Liu et al., 2021). Additional biomarkers such as carcinoembryonic antigen (CEA), cancer antigen 15-3 (CA15-3), cancer antigen 27-29 (CA27-29), tissue polypeptide antigen (TPA), and tissue polypeptide-specific antigen (TPSA), generally demonstrate low sensitivity and/or specificity, limiting their effectiveness and often resulting in delayed detection of cancer recurrence (Nagrath et al., 2011).

Metabolomic alterations associated with BC offer the potential to identify novel diagnostic biomarkers. Changes in gene expression, including pathogenic mutations in BRCA1 and BRCA2, can drive metabolic reprogramming that supports tumor initiation and progression (e.g., Privat et al., 2014). Numerous metabolomic studies have investigated plasma, serum, and urine as accessible biofluids for biomarker discovery, demonstrating their promise for the early detection of BC (e.g., Asiago et al., 2010; Lécuyer et al., 2018). Due to their non-invasive sampling, urine metabolomes are particularly attractive as a source of biomarkers. Urine has therefore been proposed as a potential source of biomarkers for various cancer types, including BC (Günther, 2015; Asiago et al., 2010). Metabolite profiling of urine samples using liquid chromatography–mass spectrometry identified 24 potential biomarkers. Receiver operating characteristic (ROC) analyses showed that the concentrations of N-(2-furoyl)glycine (area under the curve [AUC] = 0.902), histidine (AUC = 0.785), and D-tagatose (AUC = 0.732) were significantly elevated, whereas trigonellinamide (AUC = 0.827), L-galacto-2-heptulose (AUC = 0.817), and creatinine (AUC = 0.821) were markedly reduced in BC patients compared with healthy controls (Park et al., 2019). A proton nuclear magnetic resonance (¹H-NMR)–based study similarly identified ten metabolites including creatine, glycine, trimethylamine N-oxide, and serine, that differed significantly between BC patients and healthy individuals. Pathway analysis indicated notable alterations in glycine and butanoate metabolism among BC patients (Silva et al., 2019).

Despite these advances, a more precise characterization of the molecular events underlying BC development may be achieved by focusing on extracellular vesicles (EVs). EVs are lipid-bound vesicles released by cells into the extracellular environment (Bebelman et al., 2018; Zaborowski et al., 2015). The major subtypes of EVs include microvesicles (MVs), exosomes, and apoptotic bodies, which are distinguished by their biogenesis, release mechanisms, size, composition, and functional roles. EVs are currently being explored for their potential in disease diagnosis, as natural drug-delivery vehicles, and as therapeutic tools; however, they also contribute to pathological processes, including cancer progression (Bebelman et al., 2018; Bernatova et al., 2025; X. Zhang et al., 2023). EVs transfer oncogenic proteins, RNAs, and metabolites that promote tumor growth, invasion, metastasis and immune evasion. Thus, EVs reshape the tumor microenvironment and prepare pre-metastatic niches (Greening et al., 2025)

In the present study, we use direct infusion–mass spectrometry (DI-MS) to compare whole-urine and extracellular vesicle (EV)–derived metabolomes, assessing the extent to which each reflects the other and their ability to differentiate between breast cancer (BC), benign breast disease (BBD), symptom controls (SC), and healthy individuals (HC). We observed a close correlation between whole urine and EV metabolomes. Each metabolome suggested a small number of metabolite changes that were specific to BC, but these changes were common to both the whole and EV metabolomes. This could indicate that the urinary EVs describe distinctive aspects of the breast carcinogenic process.

## Materials and Methods

### Ethics approval and participant recruitment

Ethical approval for this study came from the Health Research Authority (HRA) and Health and Care Research Wales (HCRW) (IRAS Project ID: 306872; Protocol No: AU/IBERS/010; REC Reference: 21/SC/0411; CPMS Study ID: 54143).

Eligible participants were adult women (aged >18 years) presenting with breast cancer (BC)–related symptoms such as a palpable breast lump or breast pain and a suspected diagnosis of BC. Midflow urine samples were collected before standard diagnostic procedures, which included breast examination, mammography, ultrasonography, and/or needle core biopsy. Individuals were recruited during routine breast screening appointments at the rapid access of Glan Clwyd and Wrexham Maelor Hospitals (UK), between January to June 2025. Healthy female volunteers with no history of BC or other malignancies were recruited at Aberystwyth University (UK). A total of 55 urine samples were obtained from consenting adult females aged 23–91 years (mean age: 55 ± 14.88 years). Baseline demographic characteristics including factors associated with breast cancer risk were recorded for all participants (Table 1), and clinical characteristics of BC patients were listed (Table 2).

**Table 1.** Demographic characteristics of participants recruited (n=55).

| Sample groups |  | Breast cancer (BC) | Benign breast disease (BBD) | Symptom control (SC) | Healthy control (HC) | Total |
| --- | --- | --- | --- | --- | --- | --- |
| Number of participants |  | 42 | 3 | 4 | 6 | 55 |
| Age range (Median ± SD) | Overall | 31-91 (58.59±13.08) | 32-52 (41 ± 7.87) | 32-70 (51 ± 17.34) | 23-66 (43.33± 15.34) | 23-91 (55.12± 14.88) |
| Number of participants by age range, n (%) | <40 | 3 (7.14) | 2 (66.67) | 2 (50) | 3 (50) | 10 (18.18) |
|  | 41-60 | 18 (42.85) | 1 (33.34) | 0 (0) | 2 (33.34) | 21 (38.18) |
|  | 61-80 | 15 (35.71) | 0 (0) | 2 (50) | 1 (16.67) | 18 (32.73) |
|  | >81 | 2 (4.76) | 0 (0) | 0 (0) | 0 (0) | 0 (0) |
|  | Not stated | 4 | 0 (0) | 0 (0) | 0 (0) | 4 (7.27) |
| BMI (kg/m<sup>2</sup>) (Mean ± SD) |  | 26.34(±6.62) | 24.77 (±8.25) | 27.5 (±5.44) | 27.19 (±6.29) | 25.88 (±5.61) |
| Smoking habits, n (%) | Current | 6 (16.67) | 0 (0) | 0 (20.43) | 1 (5.41) | 7 (15.03) |
|  | Ex | 12 (33.34) | 0 (0) | 1 (26.88) | 1 (13.51) | 14 (29.79) |
| (n=36) | Never | 18 (50.00) | 3 (8.34) | 3 (52.69) | 4 (81.08) | 28 (55.18) |
| **Family history with BC, (%)** |  | 13 (30.95) | 2 (66.67) | 2 (50) | 2 (33.34) | 19 (34.54) |
| **Family history with other cancers (%)** |  | 11 (26.19) | 0 (0) | 3 (75) | 4 (66.67) | 18 (32.73) |

**Table 2.**
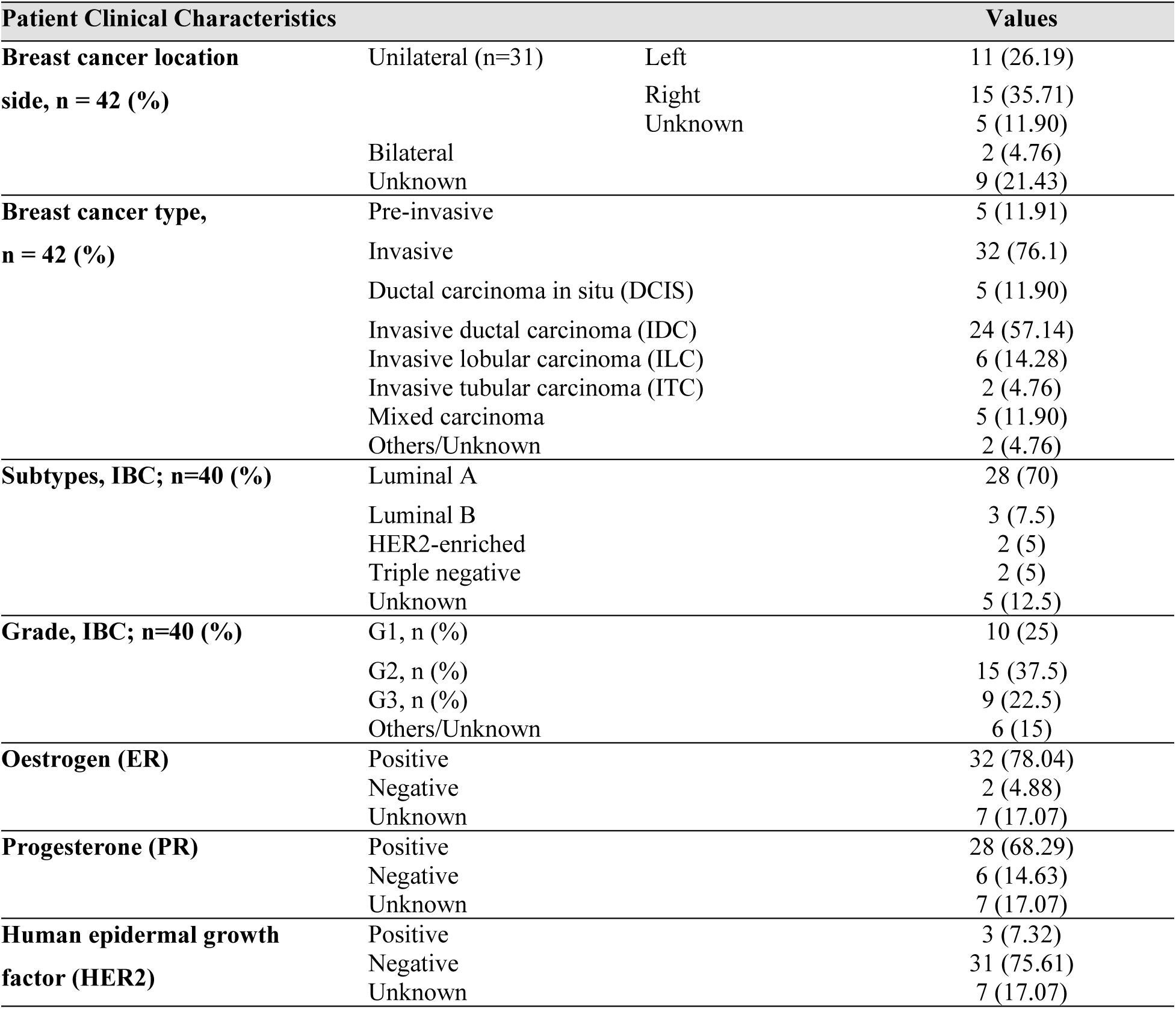
Clinical characteristics of breast cancer (BC) patients (n=42).

### Urine whole metabolomic sample preparation

The extraction of low molecular weight metabolites from urine was based on Kirwan et al. (2014), and Marques et al. (2022), with modifications, and EV-derived urine preparation and extraction was based on McGravey et al. (2024). Midstream urine samples, ranging from 10 to 40 mL, were gathered in a sterile container and immediately preserved at −80°C for long-term storage until further analysis. Aliquots of 1 mL urine sample was transferred to the microcentrifuge tubes (Greiner Bio-One, Germany) and centrifuged at 1000 x *g* for 5 minutes at 4°C. Samples were normalised to 1.006 with sterile distilled water based on specific gravity measurements using the OPTI hand-held refractometer. The samples were vortexed briefly and subjected to centrifugation at 14,000 rpm for 4 min at 4°C. A total of 160 µL of methanol/water (70:30, v/v) was added to 40 µL of adjusted urine samples and were transferred into the micro-insert of HPLC glass vials for DI-MS analysis. Aliquot of pooled samples were used as quality control (“master mixes”), whilst 200 µL of methanol/water (70:30, v/v) were used as sample blanks.

### Urine EVs Metabolomic Samples Preparation

Samples from the same cohort of urine samples were thawed and centrifuged at 365 × g for 10 min at 4 °C to remove cellular debris, and the supernatant was collected. Protease inhibitor solution (20 µL of 50× stock per 1 mL sample; Pierce, Thermo Fisher) was added. Supernatants were filtered through 0.22 µm syringe filters and concentrated using Amicon Ultra centrifugal filters (Millipore) by centrifugation at 4000 × g for 20 min at 4 °C, reducing the sample volume from 5 mL to <0.5 mL. Concentrated samples (ultrafiltrate) were recovered directly from the filter device and stored at −20 °C until further use(McGravey et al., 2024).

Urine ultrafiltrate samples were thawed on ice and centrifuged at 1,500 × g for 10 min, followed by 3,000 × g for 10 min at a temperature of 4 °C, before loading onto the Izon Automatic Fraction Collector (AFC). The size-exclusion chromatography (SEC) columns (qEV original, 70 nm or 75 nm) were equilibrated to room temperature and assembled on the AFC in accordance with the manufacturer’s guidelines(McGravey et al., 2024) . Sodium phosphate buffer (0.22 µm filtered) served as the running buffer. Following system priming, 0.5 mL of each sample was loaded onto the SEC column and eluted as per the manufacturer’s instructions. Fractions of 1.5 mL were collected; the first eluted fraction contained EVs) while the subsequent fractions (dESP1–dESP9) comprised soluble protein components. Columns were flushed between runs with sodium phosphate buffer and cleaned using 0.5 M NaOH. For storage, columns were filled with 2 mL of 20% ethanol and maintained at 4 °C. Each SEC column was used for a maximum of five separation runs.

### EVs Visualization using Transmission Electron Microscope (TEM)

Carbon/formvar-coated copper grids were placed (shiny side down) onto 10 µL of EV eluted sample spotted on an ethanol-cleaned agar surface and incubated on ice for 45 min to allow adsorption. Grids were then transferred to 10 µL of 4% (v/v) uranyl acetate for 5 min for negative staining. Excess stain was removed using filter paper, and grids were air-dried at room temperature for ≥24 hours before TEM imaging (McGravey et al., 2024).

### Metabolite profiling using direct infusion-mass spectrometry (DI-MS)

Metabolite profiling was performed using a Q Exactive mass spectrometer (Thermo Fisher Scientific, Bremen, Germany) operated in both positive and negative electrospray ionization (ESI) modes, as described previously (Marques *et al*., 2022). Samples (20 µL) were directly infused into a continuous flow of methanol: water (70:30, v/v) at 100 µL·min⁻¹. Spectra were acquired over an *m/z* range of 70–1000 using a mass resolution of 140,000 (at *m/z* 200). Data acquisition was carried out for 3.5 min with a mass accuracy of 3 (±1) ppm. Instrument source parameters were optimized according to the manufacturer’s recommendations.

Spectral binning used BinneR (Finch et al., 2022) which also eliminated anomalous single scan m/z events, the averaged of spectra across the infusion profile. The modal accurate *m/z* was then extracted for each bin spectra and combined in a single intensity matrix (runs *x m/z*) for each ion mode. The derived data are presented in Table S1. Key metabolites were tentatively identified using the HMDB (https://hmdb.ca/) and KEGG (https://www.genome.jp/kegg/) databases based on predicted masses *m/z* ratios and likely ionization forms. All isotopes/adducts were considered when deriving the identities for individual *m/z*. Metabolite identifications considering the following possible adducts: [M+]+, [M+H]+, [M+NH4]+, [M+Na]+, [M+K]+, [M-NH2+H]+, [M-CO2H+H]+, [M-H2O+H]+; [M−]−, [M−H]−, [M+Na−2H]−, [M+Cl]−, [M+K−2H]−. Correlations between multiple adducts of the suspected metabolites was an output of the BinneR programme and used in the identification process. This conforms to Metabolomics Standards Initiative (MSI) Level 2.

### Statistical Analysis

Data processing and statistical analyses used the R-based MetaboAnalyst v6.0 platform (Pang et al. 2024. The data matrix underwent log₁₀ transformation and Pareto scaling. Univariate analyses, which included ANOVA for comparisons among multiple groups and t-tests for pairwise comparisons, were conducted to assess statistically significant differences between groups. For multivariate chemometric analysis, supervised partial least squares–discriminant analysis (PLS-DA) was employed to illustrate separations based on metabolic profiles between groups. The MetaboAnalyst metabolite ID conversion tool, and *mummichog* analysis (Li et al., 2013) was utilized to identify enriched metabolic pathways (Pang et al., 2024).

### Integrative Analysis

The MixOmics package in the RStudio environment was used to compare metabolites identified from whole-urine and EV-derived urine metabolomic datasets. A design matrix representing the relationships between datasets was constructed using the minimum pairwise correlations obtained from the first component. Based on the scree plot, components explaining at least 80% of the total variance were selected as the ncomp parameter for the block.plsda the function used to generate the Circos plot within the DIABLO (Data Integration Analysis for Biomarker Discovery using Latent Variable Approaches for Omics Studies) framework. The DIABLO analysis was visualized using the plotDiablo( ) and network( ) functions to produce a matrix scatter plot and network plot, respectively. These visualizations displayed (i) the distribution of samples across latent components and (ii) the positive and negative correlations between the two metabolomic datasets (Rohart et al., 2017; Singh et al., 2016, 2019).

## Results

### Evaluation of whole urinary metabolomes

Untargeted metabolite profiling of urine samples from 55 participants were assessed through DI-MS, which detected a total of 5352 metabolic features (mass-to-charge ratio [*m/z*] values/adducts) (Table 3). To assess variation that could be linked to the experimental class partial least squares–discriminant analysis (PLS-DA) was used (Figure 1). PLS-DA indicated a clear separation of metabolite profiles between the BC/ benign breast disease (BBD) groups and the symptom control (SC), and healthy control (HC) groups. There was a separation between the BC/BBD group but considerable overlap between the HC and SC clusters (Figure 1a). Pairwise comparisons using PLS-DA highlighted differences between BC and BBD groups (Figure 1b, based on ∼9.5 % total variation) but also with SC (Figure 1c, based on ∼14.7 % total variation) and healthy (Figure 1d, based on ∼12.3 % total variation) controls.

**Figure 1.**
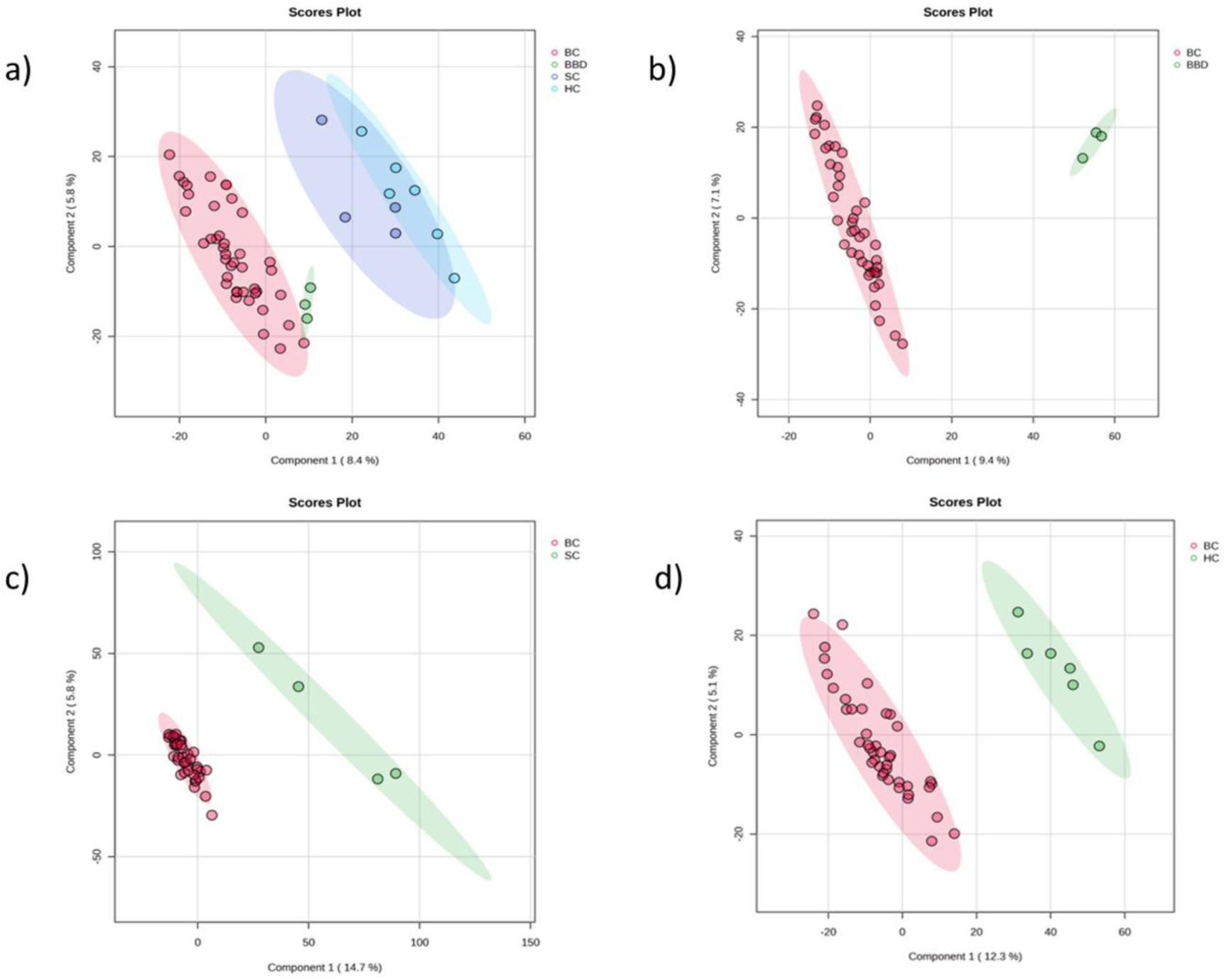
Partial Least-Squares Discriminant Analysis (PLS-DA) of whole urine metabolomes comparing the a) four study groups (ņ=55) breast cancer (BC, n=42); benign breast disease (BBD n=3); symptom control (SC n=4) and healthy control (HC, n=6). Also, pairwise PLS-DA between BC vs b) BBD, c) SC, and d) HC.

**Table 3.** The total number of ions detected (positive and negative adducts) in total urine samples and also showing significant differences (ANOVA, *P* < 0.05, corrected for false discovery rates [FDR]) when comparing breast cancer (BC) against benign breast disease (BBD), symptom controls (SC) and healthy controls (HC).

| Ionisation mode | Total number of ions detected | Number of ions with adjusted p-value $< 0.05$ |
| --- | --- | --- |
| Positive (p) | 2734 | 415 |
| Negative (n) | 2618 | 323 |
| Total | 5352 | 738 |

To identify sources of variation among the sample groups, analysis of variance (ANOVA) was performed. ANOVA revealed 738 features with statistically significant differences (p < 0.05, false discovery rate (FDR)-corrected) across the groups (Table 3), encompassing both positive and negative ionization modes. Subsequently, pairwise *t*-tests were conducted to assess differential urinary metabolite profiles between BC and BBD, SC, and HC. These comparisons identified 126, 204, and 115 significantly altered metabolites (FDR < 0.05), respectively, within which a smaller number were also targeted by volcano plots (Table 4).

**Table 4.** Pairwise comparisons between total urine metabolomes from breast cancer (BC) and benign breast disease (BBD), symptom controls (SC) and healthy controls (HC) indicating the total number of ions (positive and negative adducts) showing significant (*t*-test, *P* < 0.05, corrected for false discovery rates [FDR]) differences also those indicated using volcano plots incorporating 2 fold-changes (FC).

| BC vs | Number of ions with adjusted p-value$<0.05$ | | | Volcano plot | |
| --- | --- | --- | --- | --- | --- |
|  | Total | Positive ions | Negative ions | Up regulated | Down regulated |
| BBD | 126 | 60 | 66 | 31 | 0 |
| SC | 204 | 82 | 122 | 46 | 15 |
| HC | 115 | 49 | 66 | 30 | 3 |

The *m/z* were designed as differentially accumulating metabolites (DAMs) based on the results of the volcano plots (Table 4). The DAMs were tentatively identified and are compared using a heatmap in Figure 2a. Dilycerylphosphoryl, N-eicosapentaenoyl, 4-acetamidobutanoic acid, sphinganine-1-phosphate, and tetracosahexaenoic acid exhibited significantly higher levels in the BC group (p < 0.05) compared to the other three groups. Cholic acid, mesaconic acid, D-glucurono-6,3-lactone, glycocholic acid, and prostaglandin F2α were relatively lower in the BC group (p < 0.05). A metabolite set enrichment analysis (MSEA) was conducted to ascertain the metabolic pathways linked to the DAMs. A total of 25 pathways were enriched (with the most notable pathways being tyrosine metabolism, pentose and glucuronate interconversions, arginine and proline metabolism, and butanoate metabolism (Figure 2 b).

**Figure 2.**
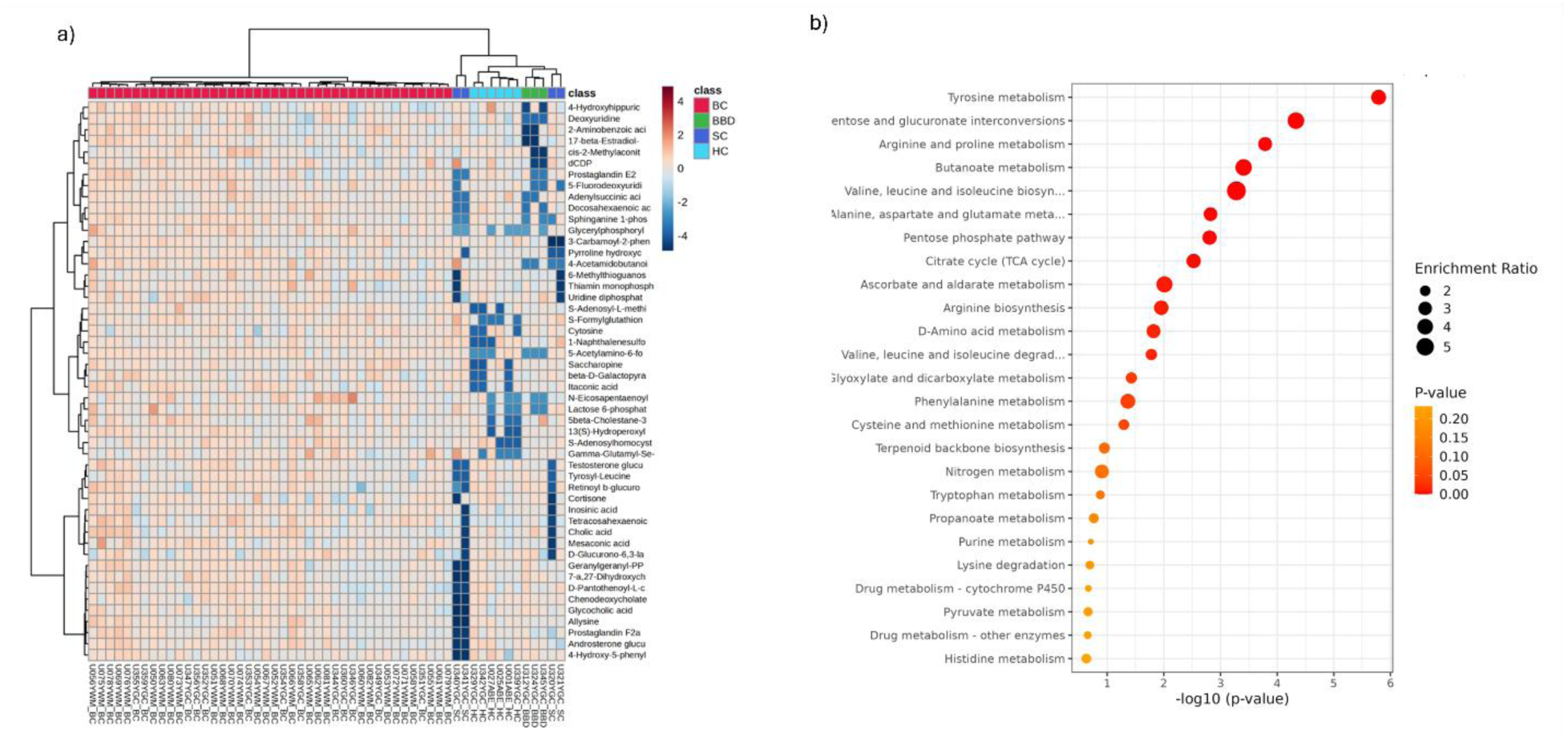
(a) Heatmap showing differential accumulating metabolites (DAMs) in whole urine metabolomes when comparing between breast cancer (BC. n=42) with benign breast disease (BBD n—3); symptom control (SC n-4) and healthy control (HC, n=6) sample groups, (b) significantly altered metabolic pathway as indicated by the DAMs. The enrichment ratio is depicted, where the size of the bubble indicates the level of enrichment.

### Extracellular Vesicles (EV)-Urine Metabolites

Purified EVs were profiled by DI-MS and a total of 6712 *m/z* features were detected (3515 positive ions and 3197 negative ions) (Table 5). PLS-DA of the derived data (Figure 3a) suggest closer similarities between the experimental classes than seen with whole urine metabolomes (Figure 1a) but BC samples appeared to be distinctive. Pairwise PLS-DA comparisons suggested that there were difference between the experimental groups; BC and BBD groups (Figure 3b, based on ∼11.5 % total variation) but also with symptom (Figure 3c, based on ∼16.8 % total variation) and healthy (Figure 3d, based on ∼29.1 % total variation) controls. ANOVA (*P* <0.05, correcting for FDR) identified 824 *m/z* as significantly differing between the classes (Table 5). Then *t*-tests (*P* <0.05, correcting for FDR) were used to identify significant pairwise comparisons. Pairwise comparisons were also assessed using volcano plots with a threshold of 2 x FC (Table 6). *M/z* targeted by both *t-*test and volcano plots were considered to be EV DAMs.

**Figure 3.**
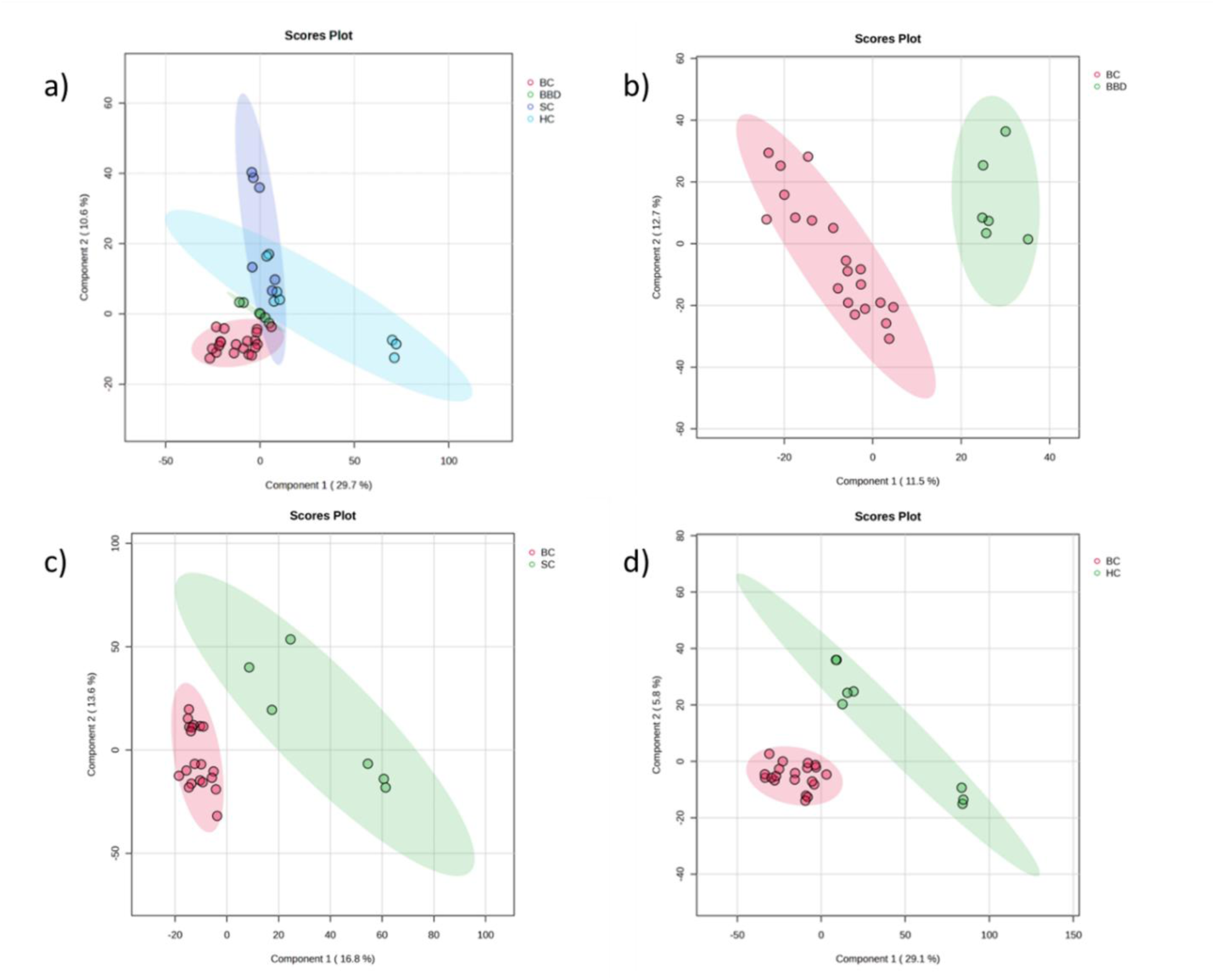
Partial Least-Squares Discriminant Analysis (PLS-DA) of urinary extracellular vesicle metabolomes comparing the a) four study groups (n=55) breast cancer (BC, n=42); benign breast disease (BBD n=3); symptom control (SC n=4) and healthy control (HC, n=6). Also, pairwise PLS-DA between BC vs b) BBD, c) SC, and d) HC.

**Table 5.** The total number of ions detected (positive and negative adducts) in extracellular vesicle (EV) urine samples and also showing significant differences (ANOVA, *P* < 0.05, corrected for false discovery rates [FDR]) when comparing breast cancer (BC), vs benign breast disease (BBD), symptom controls (SC) and healthy controls (HC).

| Ionisation mode | Total number of ions detected | Number of ions with adjusted p-value $< 0.05$ |
| --- | --- | --- |
| Positive (p) | 3515 | 326 |
| Negative (n) | 3197 | 498 |
| Total | 6712 | 824 |

**Table 6.** Pairwise comparisons between extracellular vesicle (EV) urine metabolomes of breast cancer (BC) and benign breast disease (BBD), symptom controls (SC) and healthy controls (HC) indicating the total number of ions (positive and negative adducts) showing significant (*t*-test, *P* < 0.05, corrected for false discovery rates [FDR]) differences also those indicated using volcano plots incorporating 2 fold-changes (FC).

| BC vs | Number of ions with adjusted p-value $< 0.05$ | | | Volcano plot | |
| --- | --- | --- | --- | --- | --- |
|  | Total | Positive | Negative | Up regulated | Downregulated |
| BBD | 132 | 62 | 70 | 19 | 0 |
| SC | 240 | 94 | 146 | 0 | 9 |
| HC | 111 | 45 | 66 | 23 | 0 |

DAMs from urine EV samples from the BC, BBD, SC, and HC control group comparisons were tentatively identified and illustrated using a heatmap (Figure 4a). DAMs such as tyramine, L-carnitine, histidine, and adenosine monophosphate exhibited significantly higher levels in the BC group (p < 0.05) compared to the other three groups. Coenzyme A, allysine, and proline were present at lower levels in the BC group. A MSEA approach was again used to suggest which pathway were most represented by the DAMs (Figure 4b). Approximately 25 pathways were found to be enriched with the most promient being arginine and proline metabolism, tryptophan, nitrogen, purine metabolism, and pyrimidine metabolism.

**Figure 4.**
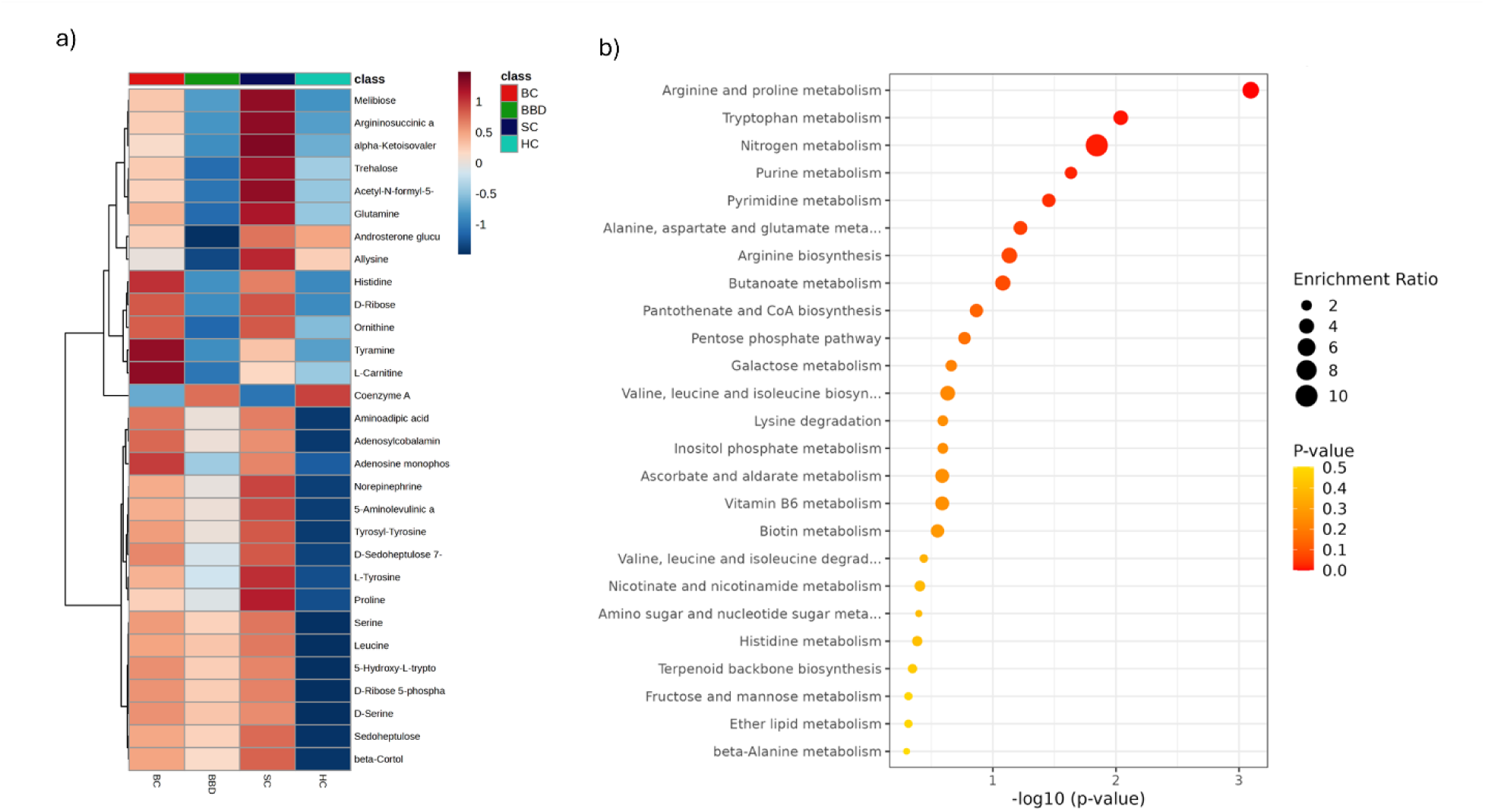
(a) Heatmap showing differential accumulating metabolites (DAMs) in urinary extracellular vesicles (EV) comparing between breast cancer (BC, n=42) with benign breast disease (BBD n=3); symptom controls (SC n=4) and healthy controls (HC, n=6) sample groups, (b) significantly altered metabolic pathway as indicated by the DAMs. The enrichment ratio is depicted, where the size of the bubble indicates the level of enrichment.

### Integrative analysis

To relate the whole urinary and EV metabolomes, the key explanatory DAMs were compared (Table 7). These were further ranked by area under the curve (AUC) values derived following Receiver Operator Characteristic (ROC) curve assessments. Many DAMs had AUC values > 0.7, considered to be the threshold for diagnostic relevance (Hosmer et al., 2013). Visual comparison of the two DAM populations suggested no overlap.

**Table 7:**
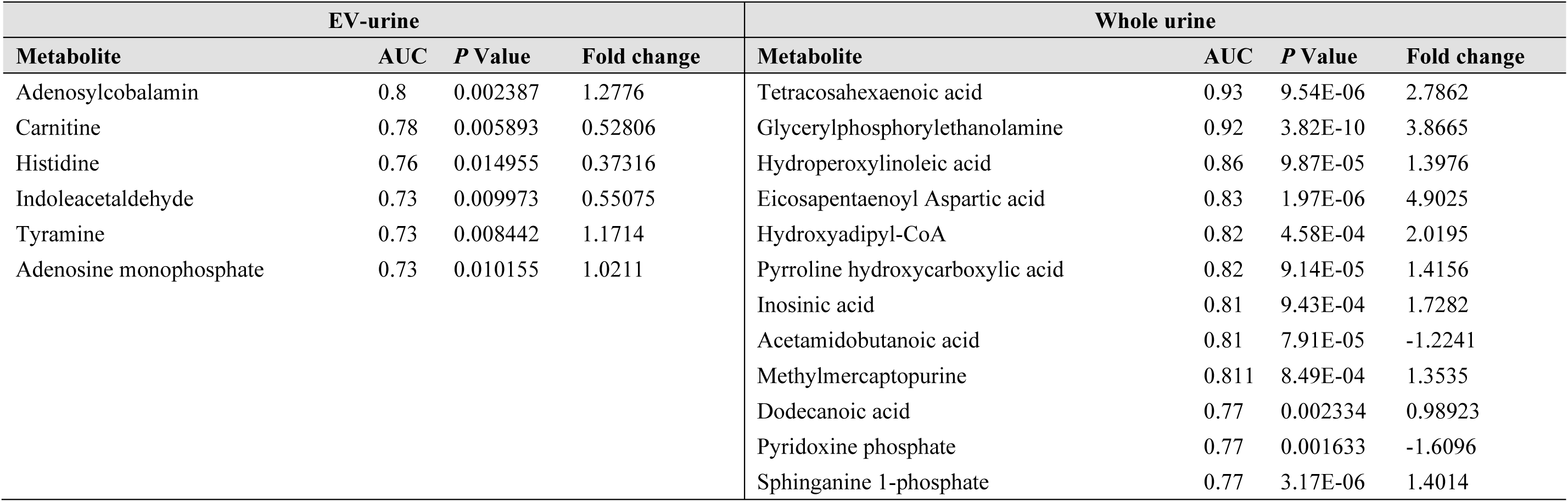
Comparison of differentially accumulating metabolites (DAMs) between breast cancer and all other experimental classes (benign breast disease [BBD]; symptom controls [SC] and healthy controls [HC]) in urine extracellular vesicle (EV) and whole urine metabolomes. Area under the curve (AUC) values, *P* values and fold changes in BC versus all other classes comparisons are shown.

Further comparisons of the whole urine_whole and urine_ev metabolomes were evaluated using the DIABLO algorithm (block PLS) from the *mixOmics* R package. Pairwise DIABLO suggested exceptionally strong linear relationship between the two data sets (Figure S1a). To investigate this further, individual metabolite with a high degree of correlation circos plot where urine_whole and urine_ev datasets (correlation cutoff of r ≥ 0.95, [Figure S1b]). These suggested that D-ribose, glutamine, aminoadipic acid, testosterone glucuronide, allysine, indoleacetaldehyde, androsterone glucuronide, and Acetyl N-formyl-5 methoxykynurenamine which were common to both urine and urine-derived EV samples.

A metabolite correlation network was constructed to highlight the relationships between metabolites present in whole urine and in urinary EVs (Figure 5). This indicated a large and interconnected module comprising pentose phosphate pathway intermediates (D-ribose, D-ribose-5-phosphate, D-sedoheptulose-7-phosphate). A second module linking α-ketoisovaleric acid, norepinephrine, and trehalose reflected coordinated amino acid catabolism. Steroid conjugates formed a distinct cluster of urine–EV metabolite pairs. Taken together, these data suggest considerable overlaps in whole urine and EV metabolomes. However, within these similar metabolomes there are distinct metabolite changes that reflect the development of BC (Table 7).

**Figure 5:**
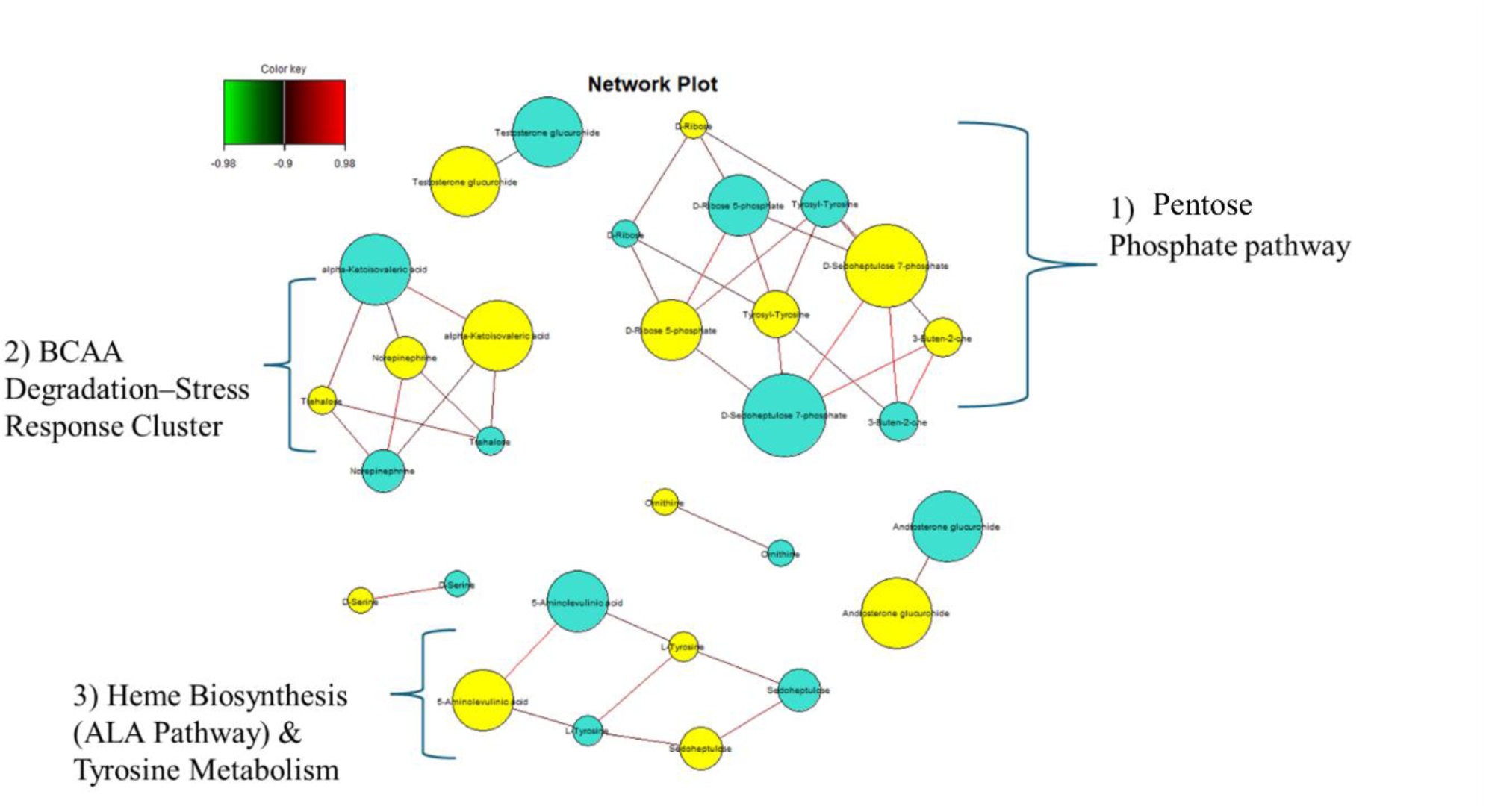
A network plot linking key features in the whole urinary and urinary extracellular vesicle (EV) metabolomes. Indicated are 1) Pentose & phosphate pathways, 2) branched chain amino acid( BCAA) degradation-stress response cluster and 3) Heme biosynthesis (ALA) and tyrosine metabolism.

## Discussion

BC is a highly heterogeneous condition characterized by multiple subtypes with unique morphologies and clinical implications, resulting in varying response patterns to different treatment approaches and clinical outcomes (Carvalho et al., 2025). This heterogeneity poses a challenge to the diagnosis and treatment of BC. In recent years, metabolomics has emerged as an important tool in cancer research, paving the way for new opportunities in biomarker discovery, disease stratification, and targeted therapies (Yang et al., 2020).

Metabolomic analyses of plasma, serum, saliva, and tissue from BC patients have provided key insights into disease-associated metabolic alterations (Jiang et al., 2025; More et al., 2017; Wu et al., 2024). However, the clinical utility of such markers is limited due to the invasive nature of sample collection and the requirement for technically demanding, high-cost analytical procedures (Hart et al., 2016). In contrast, urine offers a practical, non-invasive, and cost-effective alternative for monitoring metabolic changes. Although, a waste product, urine provides a rich source of physiological information and reflects systemic metabolic fluctuations due to its dynamic relationship with blood homeostasis (Bouatra et al., 2013). Its ease of collection, storage, and processing, combined with patient comfort and suitability for repeated sampling, makes urine a valid alternative to tissue, plasma, or serum sampling (Emwas et al., 2015; A. Zhang et al., 2012). Several studies have reported the use of urine as a valuable source for metabolite detection in BC patients (Park et al., 2019; Yang et al., 2023; Zahran et al., 2021). Key metabolite shifts with BC included amino acids, organic acids, and nucleosides as putative biomarkers (Chen et al., 2016), homovanillate, 4-hydroxyphenylacetate, 5-hydroxyindoleacetate, and urea as metabolites (Nam et al., 2009).

In this study, DI-MS was used to characterize both whole-urine and EV–derived urine (urine_EV) metabolomes in BC compared to BBD, SC and HC classes. We observed that BC was associated with distinct urinary metabolic signatures, detectable both in whole urine and in EV-enriched fractions. Interestingly, although there was considerable continuity between the two metabolomes (Figure S1), the key changes linked to BC, were distinctive (Table 7). The metabolic correlation from both the Circos plot and the network plot highlighted the similarity between metabolites freely circulating in urine and those encapsulated within urinary EVs (Figure S1). However, the differences in the clinical classes, support the concept of selective EV cargo loading, likely reflecting active metabolic processes occurring in renal epithelial cells or in circulating tumor-derived vesicles.

### Whole-Urine Metabolome in BC

Untargeted profiling of whole urine targeted several metabolites that increased with BC compared to other classes and had AUC > 0.7, suggestive of diagnostic potential (Table 7). These included glyceryl phosphoryl derivatives, N-eicosapentaenoyl species, sphinganine-1-phosphate (S1P), and long-chain polyunsaturated fatty acids such as tetracosahexaenoic acid); suggestive of enhanced lipid membrane biosynthesis and sphingolipid signaling. The enrichment of S1P aligns with its established role in supporting BC proliferation, angiogenesis, and metastatic dissemination through S1P-receptor–mediated pathways (Pyne & Pyne, 2020;Pyne et al., 2016; Corsetto et al., 2023). Tetracosahexaenoic acid had the highest AUC value at 0.924. This metabolite is not been previously linked to BC but has been suggested as a potential biomarker for early detection of cervical cancer (Yang et al., 2017). Metabolomic studies of cervical cancer and esophageal squamous cell carcinoma suggest tetracosahexaenoic acid changes could be indicative of dysregulated lipid metabolism. In contrast, lower amounts of cholic acid, glycocholic acid, mesaconic acid, D-glucurono-6,3-lactone, and prostaglandin-F2α in BC suggest a disruption in liver detoxification, sterol metabolism, and the turnover of inflammatory lipids, processes that are known to be affected in cancer due to metabolic–endocrine interactions driven by tumors (Brodie & Sabnis, 2019). The decrease in glucuronide-related metabolites may point to changes in UDP-glucuronosyltransferase activity or deconjugation linked to the microbiome, which aligns with new evidence showing that tumor-related dysbiosis affects systemic metabolite levels (Zhang et al., 2020).

Pathway enrichment analysis of whole urine metabolomic profiles revealed significant alterations in several metabolic pathways, including tyrosine metabolism, pentose and glucuronate interconversions, arginine and proline metabolism, and butanoate metabolism. These pathways are closely associated with energy homeostasis, amino acid turnover, and cellular oxidative stress responses. BC cells can sustain growth under amino-acid deprivation by degrading and internalizing components of the extracellular matrix (ECM), a process not observed in non-cancerous breast cells (Nazemi et al., 2024). Tyrosine catabolism produces fumarate, which feeds into the tricarboxylic acid (TCA) cycle, thereby supporting energy production and enabling continued proliferation and migration even in nutrient-limited conditions (Nazemi et al., 2024).

### EV-Derived Urine Metabolome

Unlike whole urine, EV-derived profiles showed more distinct separation between BC and all comparator groups, with minimal overlap on PLS-DA plots (Figure 3a). This suggests that EV enrichment could enhance the detection of tumor specific.

Several metabolites, including tyramine, histidine, carnitine, and adenosine monophosphate, were elevated in BC-derived EVs. Park et al., (2019) reported increased levels of histidine in whole urine of BC patients compared to healthy individuals, with an AUC=0.799. Histidine plays several critical roles in BC biology. As the precursor of histamine, histidine supports elevated histamine production in tumors, driven by increased histidine decarboxylase (HDC) activity, which promotes inflammation and BC cell proliferation(Azimi et al., 2024) Histamine also modulates the tumor immune microenvironment, contributing to T-cell dysfunction and reduced responsiveness to immunotherapy (Azimi et al., 2024). Moreover, HDC expression correlates with clinicopathological features, suggesting a potential prognostic value (Azimi et al., 2024). Circulating histidine levels may further predict treatment-related toxicities, such as paclitaxel-induced peripheral neuropathy, highlighting its relevance for personalized therapy (Sun et al., 2018)

In our study, carnitine levels were also elevated in BC patients compared with the other groups (AUC = 0.78; Table 7). Although metabolite-based investigations of EV-derived urine in BC remain limited, our findings are consistent with those of Sun et al. (2020), who reported significantly higher L-carnitine concentrations in BC tissue (n=170) relative to normal tissue (n=128), with strong diagnostic performance (AUC = 0.98, sensitivity 0.95, specificity 0.97) (Sun et al., 2020). Furthermore, other studies have proposed carnitine as a potential prognostic biomarker in other tumor types, including sarcoma (Lou et al., 2017). Clos-Garcia et al. reported elevated carnitine levels in urinary EVs from prostate cancer patients, indicating altered fatty acid metabolism (Clos-Garcia et al., 2018). Puhka et al. suggested that fluctuations in carnitine content within prostate cancer–derived EVs reflect a metabolic shift toward enhanced fatty acid β-oxidation (Puhka et al., 2017).

We observed that adenosine monophosphate was elevated in BC patients with AUC=0.732. This could reflect altered adenosine processing as reported by Tadokoro et al., (2020). These authors observed extracellular leakage of adenosine form BC–derived EVs. This EV-derived adenosine then binds to adenosine receptors and suppresses immune activity by inhibiting perforin release from cytotoxic T cells. Thus, elevated adenosine levels can promote immune escape, and support tumor progression. Detecting increased adenosine in EVs may therefore provide a non-invasive biomarker for BC and reflect tumor-specific metabolic alterations with high specificity.

Coenzyme A, allysine, and proline were reduced in BC urine_EV metabolites.samples. Depletion of coenzyme A–related intermediates may reflect increased tumor consumption for lipid synthesis and mitochondrial metabolism. The decreased levels of proline and allysine are consistent with aberrant collagen turnover and extracellular matrix remodeling, hallmarks of tumor invasion and metastasis.

## Conclusion

Urinary metabolomics, particularly using EV–enriched fractions, has been shown to reveals distinct BC–associated metabolic signatures that reflect tumor-specific reprogramming in lipid, amino acid, nucleotide, and oxidative stress pathways. EV-derived metabolites, including histidine, L-carnitine, and adenosine, demonstrate superior sensitivity in capturing disease-related changes compared with whole urine, highlighting their potential as non-invasive biomarkers for early detection and disease stratification. These findings underscore the promise of urine-EV metabolomics as a clinically feasible tool for breast cancer diagnostics, prognosis, and precision medicine, warranting further validation in larger cohorts.

## Data Availability

All data produced in the present study are available upon reasonable request to the authors

## ACKNOWLEDGEMENTS

N.A.A.Z was supported by a Graduate Excellence Programme (GrEP), Majlis Amanah Rakyat Malaysia (MARA) fellowship (Grant No. 330408376064). L.A.J.M is partially supported by Shandong Province “Double-Hundred Talent Plan” Teams (Grant No. WSR2023049).

---

**Figure S1.**
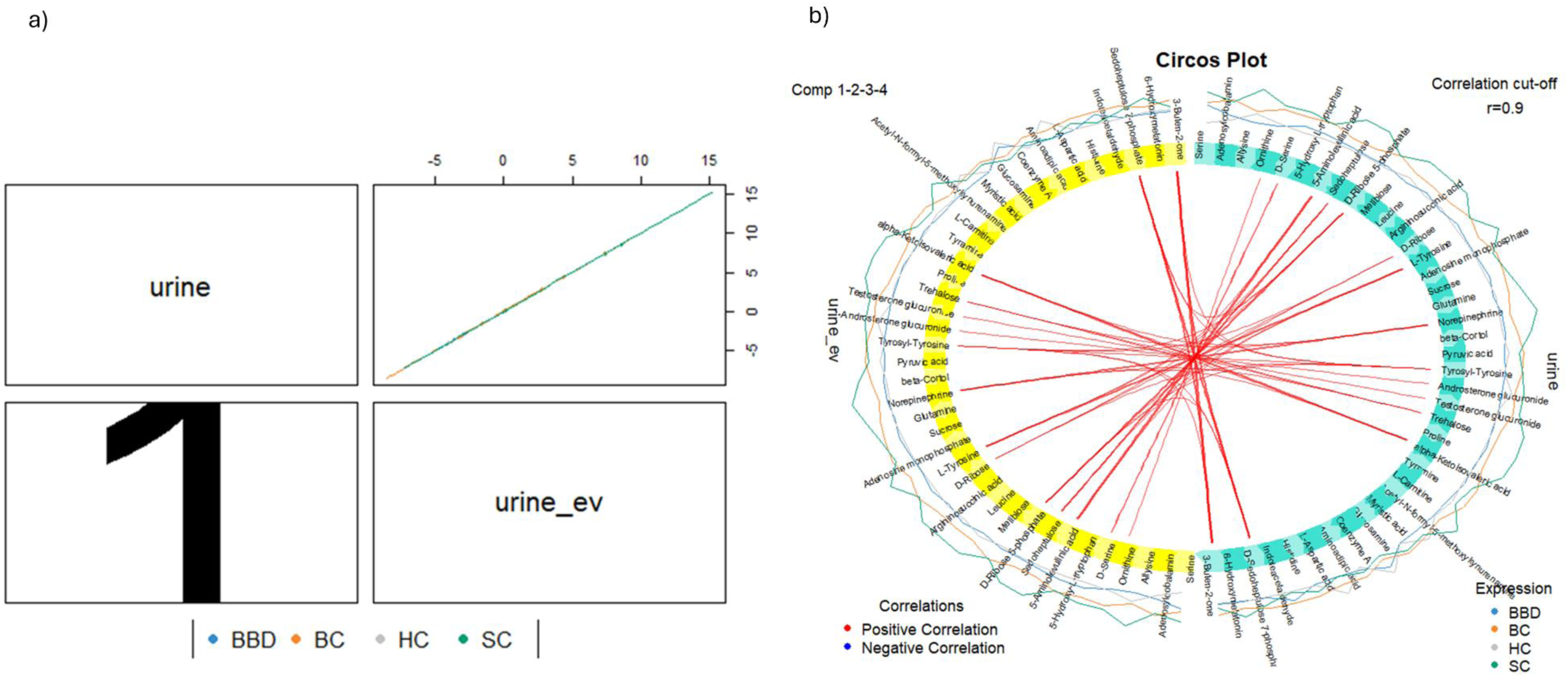
Comparing the whole urine and urinary EV metabolomes **a)** Scatter plot correlating urine_whole and urine_ev metabolomes by using the R mixomics platform. b) Circos plot linking key metabolites of whole urine( blue) and EV-urine (yellow) metabolomes when using a correlation cutoff of r ≥ 0.9, No negative correlations are observed at this threshold. The outer rings display metabolite levels for the four sample groups-BED (blue), BC (orange), HC (gtey), and SC (green)-showing group-specific variation in abundance for each metabolite.

